# Space-time conditional autoregressive modeling to estimate neighborhood-level risks for dengue fever in Cali, Colombia

**DOI:** 10.1101/2020.06.20.20136226

**Authors:** M.R. Desjardins, M.D. Eastin, R. Paul, I. Casas, E.M. Delmelle

## Abstract

Vector-borne diseases (VBDs) affect more than 1 billion people a year worldwide, cause over 1 million deaths, and cost hundreds of billions of dollars in societal costs. Mosquitoes are the most common vectors, responsible for transmitting a variety of arboviruses. Dengue fever (DENF) has been responsible for nearly 400 million infections annually. Dengue fever is primarily transmitted by female *Aedes aegypti* and *Aedes albopictus* mosquitoes. Since both *Aedes* species are peri-domestic and container-breeding mosquitoes, dengue surveillance should begin at the local level - where a variety of local factors may increase the risk of transmission. Dengue has been endemic in Colombia for decades and is notably hyperendemic in the city of Cali. For this study, we use weekly cases of DENF in Cali, Colombia from 2015-2016; and develop space-time conditional autoregressive models to quantify how DENF risk is influenced by socioeconomic, environmental, and accessibility risk factors, and lagged weather variables. Our models identify high-risk neighborhoods for DENF throughout Cali. Statistical inference is drawn under Bayesian paradigm using Markov Chain Monte Carlo techniques. The results provide detailed insight about the spatial heterogeneity of DENF risk and the associated risk factors (such as weather, proximity to *Aedes* habitats, and socioeconomic classification) at a fine-level, informing public health officials to motivate at-risk neighborhoods to take an active role in vector surveillance and control, and improving educational and surveillance resources throughout the city of Cali.

## 1. Introduction

Vector-borne diseases, more specifically mosquito-borne arboviruses are responsible for 1 billion infectious disease cases each year, globally^1,2^. Mosquitoes transmit a variety of arboviruses and are the most common vectors. Dengue fever (DENF) is a mosquito-borne disease that is responsible for the majority of the global burden of arboviruses^3^. Over 40% of humans are at risk of transmission, with incidence rising 30-fold in the last 50 years; and it is estimated that there are approximately 390 million DENF infections annually^4^. DENF is primarily transmitted by the *Aedes aegypti* and *Aedes albopictus* mosquitoes^5,6^. Both species are container-breeding mosquitoes that have become prolific in urban areas due to the widespread availability of breeding habitats^7^.

Dengue is a flavivirus that causes DENF and there are four serotypes that follow the human cycle^8^. The incubation period ranges from 3-14 days after being bit by an infected mosquito, and symptoms can last from 2-7 days^9^, however, approximately 80% of infected individuals are asymptomatic. Infection from one serotype will result in lifelong immunity to that serotype, however, secondary infection with another serotype can lead to severe forms of DENF^10^, such as dengue hemorrhagic fever (DHF) and dengue shock syndrome (DSS). DHF and DSS primarily affects pediatric patients, but it has also been found among adults (especially the elderly); and mortality from dengue is highest among children and those who have experienced DSS^11^.

It is critical to implement surveillance strategies that can improve the understanding of DENF transmission. Improving DENF surveillance can facilitate the timely reporting of disease cases, reduce underreporting, inform policymakers, increase disease awareness, define funding and research priorities^12^; ultimately reducing the economic and public health burden in at-risk locations around the world^13^. Approaches and advances in geographic information science (GIScience) and spatial epidemiology play critical roles in DENF surveillance – such as tracking diffusion and cyclic patterns, detecting clusters and mapping disease rates and risk^14^ and understanding the place-based determinants of disease transmission^15^. DENF risks and rates will vary by place and covariate data are needed to identify significant variables responsible for observable spatial patterns^16^.

Therefore, it is critical to examine the social, economic, environmental, biological, and institutional factors that may affect DENF prevalence in a particular area. Urban regions are highly complex, and neighborhoods are the scale that public health departments most effectively operate^17^. Therefore, more small-area studies in spatial epidemiology are required to effectively uncover the spatial and temporal heterogeneity of DENF rates across urban landscapes at these fine levels of granularity. For example, education, income, age, access to care, and quality of prevention strategies are known to strongly influence an individual’s susceptibility to VBDs^18,19^. Likewise, the dynamics of how temperature, precipitation, and humidity affect vector abundance and DENF transmission are critical to developing and implementing effective sub-seasonal risk model^20^ and long-term mitigation in response to climate change^21^.

Conditional autoregressive (CAR) models can be utilized to examine how DENF risk is influenced by socioeconomic, environmental, and accessibility risk factors, and lagged weather variables. For example, a particular location may be influenced by DENF rates and a variety of explanatory variables (e.g. socioeconomic status, proximity to *Aedes* habitats, etc.) contained in surrounding locations (spatial spillover/diffusion effects). CAR models can be fitted to data under Bayesian paradigm (i.e. relying on prior beliefs/borrowing information to inform future estimations) using – Bayesian hierarchical models (BHM), which are widely used techniques in geography and public health to model spatial and spatiotemporal data^22^. In short, BHMs can model complicated spatial and space-time processes by conditionally modeling the variations in data, the process, and unknown parameter^23^. The temporal extension – ST-CAR can estimate the value of a variable (e.g. disease rates) at a particular location and time, which will be related to current and past values of the surrounding locations and time periods; essentially testing for spatiotemporal dependence. ST-CAR models have been used to study the effect of air pollution on human health^24^, substance abuse and its relationship with child abuse^25^, influenza^26^, and DENF27,28,29.

More research is necessary to examine the local variations in DENF transmission dynamics at very fine spatial and temporal scales. Delmelle et al. (2016)^30^ used a geographically weighted regression (GWR^31^) model which identified six significant socioeconomic and environmental independent variables (including proximity to tire shops and population density) of DENF rates in Cali, Colombia at the neighborhood-level. However, the explanatory power of GWR and its temporal extension – GTWR^32^), is limited. Both CAR and ST-CAR models produce model-based estimates and inference derived from varying effects via spatial random fields – e.g. borrowing strength from neighborhood spatial and temporal proximity; while GWR models allow the covariates to vary in space (and time in the case of GTWR), but inference is ad hoc. In other words, ST-CAR models can estimate spatially and temporally varying associations between the dependent (e.g. disease rates) and independent variables based on locally weighted regressions in both geographic and attribute space; while GTWR can only produce local estimates in geographic space. It is therefore worthwhile to utilize ST-CAR modeling in small-area DENF studies at fine temporal scales.

This study utilizes a ST-CAR modeling approach to examine the influence of socioeconomic, environmental, weather and climate variables on DENF outbreaks in Cali, Colombia at the neighborhood- and weekly levels between 2015 and 2016. The approach can determine if DENF rates and covariates in one neighborhood are influenced by rates and covariates in surrounding neighborhoods and time periods. We also estimate disease risks using temporally lagged weather variables. Our modeling approach is capable of identifying regions with high risk clusters at the neighborhood-level. The results provide detailed insight about the spatial heterogeneity of disease risk and the associated risk factors at a fine-level; informing public health officials to motivate at-risk neighborhoods to take an active role in vector surveillance and control, and improving educational and surveillance resources throughout the city of Cali.

The remainder of this paper is as follows: section 2 provides information about Cali, the DENF cases, and candidate independent variables, the technique to select the lagged weather variables, and the ST-CAR modeling approach. Section 3 provides the modeling results, including maps of the estimated DENF rates by week for each neighborhood in Cali. Section 4 discusses key findings, strengths, limitations, and avenues of future research; and section 5 summarizes findings with concluding remarks.

## 2. Data & Methods

### 2.1 Study Area & Data

Cali is the second-largest city in Colombia and third most populous with an estimated 2010 population of 2.3 million (average density of 4,000 km^2^). The city is comprised of 340 neighborhoods (Spanish - barrios), which are classified by socioeconomic stratum (ranging from 1-6); where a ranking of 1-2 is low, 3-4 is middle, and 5-6 is high. The classifications are defined by the external physical characteristics of the dwelling, its immediate surroundings, and its urban context. For example, urban context includes variables such as poverty, social deviation, urban decay, industry, commercial; immediate surroundings include access roads, sidewalks; and the characteristics of the dwelling include front lawn, garage, façade material, door material, front of the house dimensions, and windows (income is not considered). This stratification is only applied to residential constructions^33^. Figure 1 provides a map of neighborhoods in Cali and their corresponding ranking. The average size of neighborhoods in Cali is 0.35 square kilometers. Some of the smaller neighborhoods are in the city core, which is where the city was founded. The largest neighborhood is to the south and corresponds to newer developments and is an area that houses three of the largest universities in Cali.

**Figure 1.**
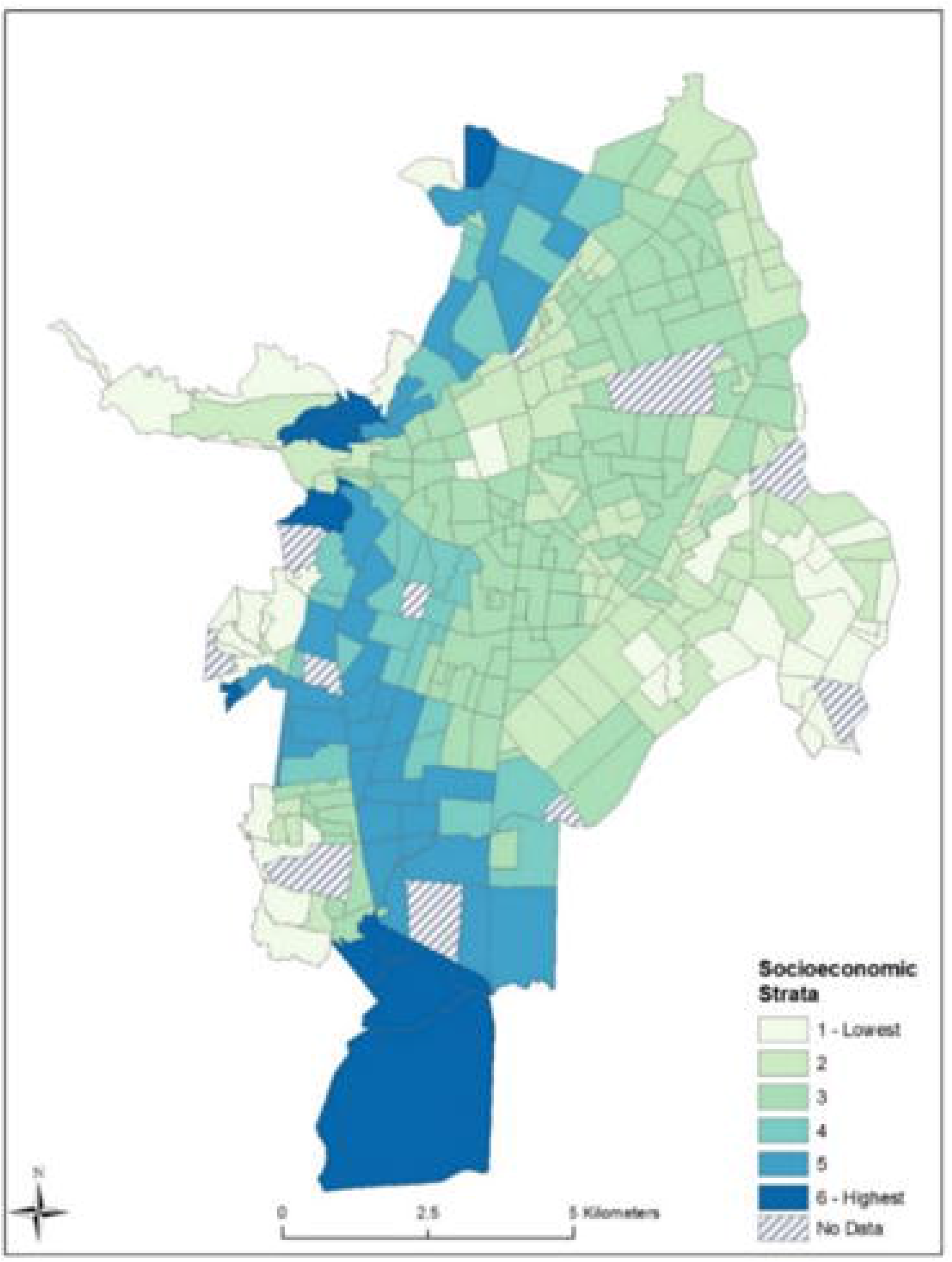
Neighborhoods in Cali, Colombia and their ranking by socioeconomic strata (1-6).

Individual cases of DENF for the years of 2015 and 2016 at the weekly level were used (Figure 2 - top), which were provided by Colombia’s National Institute of Health. Between 2015 and 2016, Cali experienced three major outbreaks: March to mid-May 2015; February to early-April 2016; and mid-June 2016 to early August 2016 (represented by the peaks in Figure 2 - top). The cases were geocoded to the neighborhood level using each neighborhood’s name as the address locator in the geocoder algorithm in ArcGIS 10.6 (ESRI, Redlands, CA) Each DENF case record contained a neighborhood where the infected individual lived (individual addresses were not available), then the geocoder aggregated the cases to a particular neighborhood after a successful match. As a result, 26,503 out of 35,498 DENF cases (74.6%) were successfully geocoded and aggregated to the neighborhood-level in Cali. Cases that were not geocoded did not have an address nor a neighborhood, therefore, it was impossible to assign coordinates to the unmatched cases.

**Figure 2.**
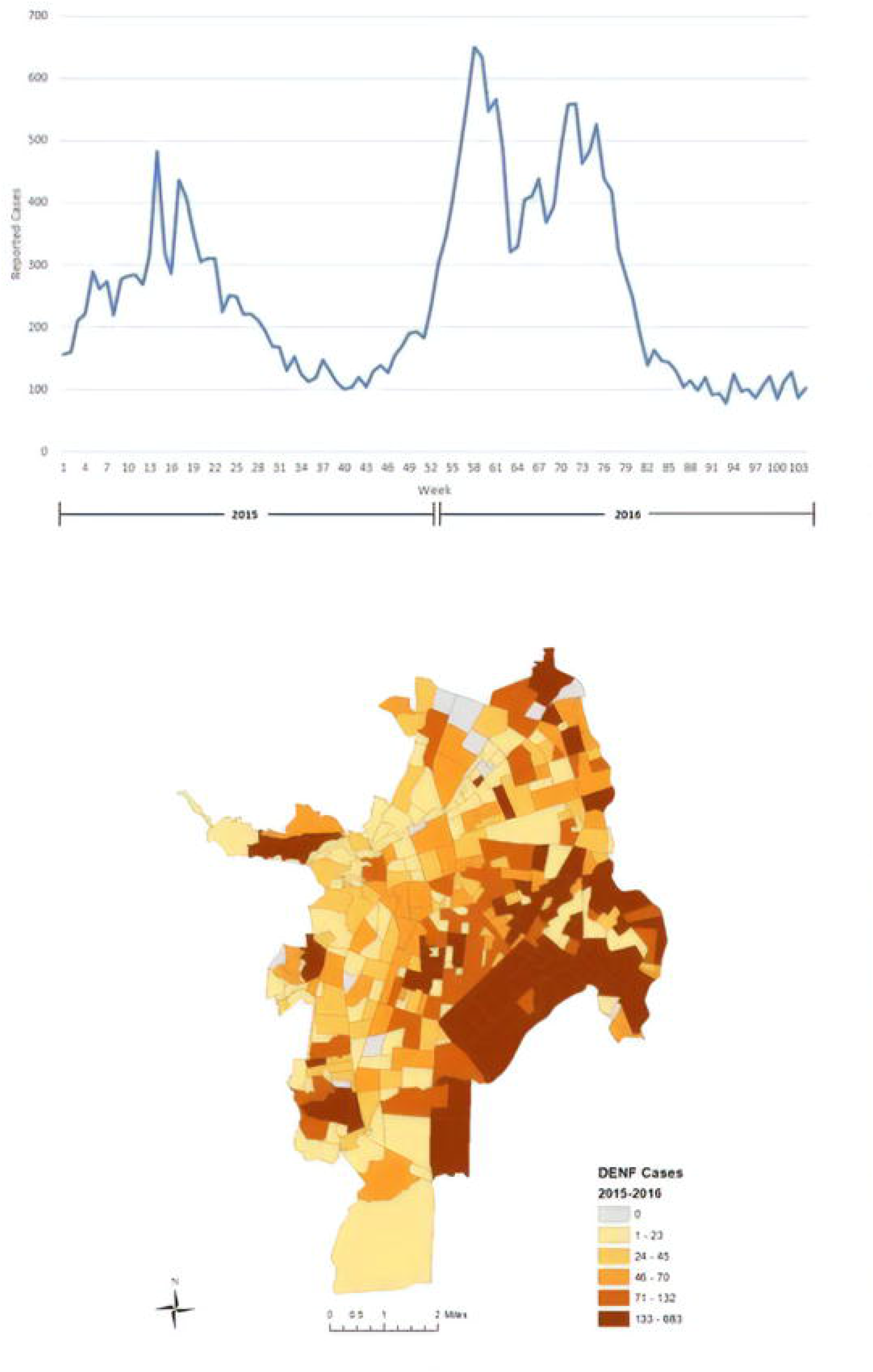
Temporal distribution of weekly DENF cases in Cali from January 2015 through December 2016 (top); spatial distribution of DENF rates per 1,000 for the study period (bottom).

Figure 2 (bottom) provides a map of the total DENF cases per neighborhood between 2015 and 2016 in Cali. The eastern portion of Cali observed the highest number of DENF cases between the two years in our study period. These neighborhoods were majority low strata (1 and 2); however, there are numerous middle strata (3 and 4) neighborhoods in the central and southern portions of Cali with a high number of DENF cases. There are also high strata neighborhoods with a high proportion of cases in the central and western regions of Cali, especially those that are adjacent to lower strata neighborhoods.

The socioeconomic and demographic data were provided by the Colombian census (either 2005 or 2010 estimates provided by the City of Cali), including population density, age, race, households with sewer and water access, educational attainment, employment status, socioeconomic stratum, among others. The last national census occurred in 2005, while the new 2018 census has yet to be released. The location of healthcare centers and the environmental variables were provided by the city of Cali (2010 data) – green zones, rivers, tire shops, water pumps, cemeteries, and plant nurseries, which were geocoded as point layers with the exception of green zones (area - polygons) and rivers (lines). The environmental variables are included as potential *Aedes* habitats. For green zones, the area of the green zones for each neighborhood was computed in square-kilometers. Similar to Delmelle et al. (2016)^30^, relative proximity to rivers, tire shops, water pumps, cemeteries, plant nurseries, and healthcare centers was computed by using kernel density estimation (KDE) – representing the density of points for each layer. KDE was also computed to produce the density of trees. Zonal statistics in ArcGIS 10.6 was used to summarize the average KDE for each neighborhood in Cali.

Finally, the weather variables selected for evaluation (Table 1) are consistent with current understanding of how weather conditions impact *Aedes* survival, abundance, and behavior, including viral transmission rates^34,20^ (Table 5). Given our goal of developing a risk model using 2015-16 weekly-level DENF data, the weather variables entail weekly summaries of 2014-16 daily meteorological observations collected at the Cali international airport and obtained from the Global Historical Climate Network archive^35^ maintained by the National Centers for Environmental Information (http://www.ncdc.noaa.gov). All weekly weather variables were computed following the methods outlined in Eastin et al. (2014)^20^, and summary statistics for the 2014-16 period are provided in Table 1. A total of 49 candidate predictor variables of DENF were evaluated in this study (Table 1). Due to the differences in units of measurements, each variable was normalized between 0 and 1 (i.e., between the maximum and minimum values listed in Table 1) for all subsequent analyses.

**Table 1:** Descriptive Statistics of the Candidate Independent Variables for Cali, Colombia (# *Mean; **Median; Total***)

| Variable ID | Variable Name | Year | Source | Value# | Range |
| --- | --- | --- | --- | --- | --- |
| **Environmental/Aedes Habitats** |  |  |  |  |  |
| Green | Area of green zones (km<sup>2</sup>) | 2010 | City of Cali | 196.4*** | 0-34.9 |
| Rivers | Relative proximity to river | 2010 | City of Cali | 1082.0 | 0-9,3539.3 |
| Tires | Relative proximity to tire shops | 2010 | City of Cali | 5.1* | 0-28.6 |
| WPumps | Relative proximity to water pumps | 2010 | City of Cali | 0.1* | 0-0.5 |
| Cemet | Relative proximity to cemeteries | 2010 | City of Cali | 2.1* | 0-9.8 |
| PNurseries | Relative proximity to plant nurseries | 2010 | City of Cali | 0.5* | 0-1.9 |
| Trees | Density of trees (km<sup>2</sup>) | 2010 | City of Cali | 1,698.9*** | 8.6-6,355.6 |
| **Healthcare Accessibility** |  |  |  |  |  |
| DistHealth | Relative proximity to a healthcare center | 2010 | City of Cali | 4.4* | 0-9.2 |
| HealthAvg | Mean healthcare center density (km<sup>2</sup>) | 2010 | City of Cali | 4.2* | 0-74.4 |
| **Socioeconomic & Demographic** |  |  |  |  |  |
| Strata | Neighborhood Stratum | 2010 | Census | 3** | 1-6 |
| Popdens | Population density (km<sup>2</sup>) | 2005 | Census | 22,099* | 0-56,814 |
| %OHH | Density of occupied households (km<sup>2</sup>) | 2005 | Census | 837.2* | 0-14,885 |
| %UHH | Density of unoccupied households (km<sup>2</sup>) | 2005 | Census | 98.6* | 0-688.4 |
| %Sew | Households with sewer (%) | 2005 | Census | 89.8* | 0-100 |
| %Water | Households with water (%) | 2005 | Census | 90.2* | 0-100 |
| %A04 | Individual age 0-4 years old (%) | 2005 | Census | 7.3* | 0-15.7 |
| %A514 | Individual age 5-14 years old (%) | 2005 | Census | 17.2* | 0-27.8 |
| %A1524 | Individual age 15-24 years old (%) | 2005 | Census | 17.5* | 0-69.2 |
| %A2539 | Individual age 25-39 years old (%) | 2005 | Census | 23.4* | 0-39.1 |
| %A4064 | Individual age 40-64 years old (%) | 2005 | Census | 26.3* | 0-47.8 |
| %A65 | Individual age 65 years old or more (%) | 2005 | Census | 8.3* | 0-23.4 |
| %Fem | Female population (%) | 2005 | Census | 53.4* | 0-100 |
| %White | White population (%) | 2005 | Census | 77.2* | 0-96.7 |
| %Black | Black population (%) | 2005 | Census | 22.1* | 0-70.6 |
| %Indig | Indigenous population (%) | 2005 | Census | 0.5* | 0-8.7 |
| %Disabled | Individuals with disabilities (%) | 2005 | Census | 1.1* | 0-5.7 |
| %NoRW | Individuals who cannot read/write (%) | 2005 | Census | 6.8* | 0-5.7 |
| %NoEduc | Individuals with no education (%) | 2005 | Census | 3.6* | 0-16.8 |
| %LowEduc | Individuals with low education (%) | 2005 | Census | 26.9* | 0-52.8 |
| %MedEduc | Individuals with medium education (%) | 2005 | Census | 49.6* | 0-71.3 |
| %HighEduc | Individuals with high education (%) | 2005 | Census | 19.9* | 0-56.0 |
| %Work | Employed individuals (%) | 2005 | Census | 41.1* | 0-58.9 |
| %Unem | Unemployed individuals (%) | 2005 | Census | 1.5* | 0-10.3 |
| %Retired | Retired individuals (%) | 2005 | Census | 3.9* | 0-15.9 |
| %HW | Individuals doing housework (%) | 2005 | Census | 13.6* | 0-18 |
| %Students | Students (%) | 2005 | Census | 22.9* | 0-36.8 |
| %Married | Married individuals (%) | 2005 | Census | 19.0* | 0-37.9 |
| %Single | Single individuals (%) | 2005 | Census | 38.9* | 0-100 |
| **Weather (Weekly Observations)** |  |  |  |  |  |
| Tavg | Mean Temperature of (°C) | 2014-2016 | City of Cali | 24.2* | 22.5-25.7 |
| Tmax | Mean Maximum Temperature (°C) | 2014-2016 | City of Cali | 31.2* | 28.5-34.2 |
| Tmin | Mean Minimum Temperature (°C) | 2014-2016 | City of Cali | 19.4* | 17.2-20.7 |
| DTRavg | Mean Daily Temperature Range (°C) | 2014-2016 | City of Cali | 11.8* | 9-15.7 |
| DTRmax | Maximum Daily Temperature Range (°C) | 2014-2016 | City of Cali | 14.3* | 10.6-21 |
| RHavg | Mean Relative Humidity (%) | 2014-2016 | City of Cali | 73.3* | 63.9-83.7 |
| RHrng | Relative Humidity Range (%) | 2014-2016 | City of Cali | 10.6* | 2.7-24.1 |
| RainT | Total Rain (mm) | 2014-2016 | City of Cali | 12.3* | 0-103 |
| RainD | Total Days with Measurable Rainfall | 2014-2016 | City of Cali | 2.8* | 0-7 |
| CoolD | Days with Minimum Temperature < 18 (°C) | 2014-2016 | City of Cali | 0.6* | 0-5 |
| WarmD | Days with Maximum Temperature > 32 (°C) | 2014-2016 | City of Cali | 2.0* | 0-7 |

### 2.2 Methodology

#### 2.2.1 Principal component analysis & VIF testing

Due to the large number of socioeconomic, demographic variables, and environmental variables (n = 36), we utilized Variance Inflation Factor (VIF)^36^ testing and a principal component analysis (PCA^37^) in Stata. We did not include all variables in the PCA because we wanted to interpret particular independent variables of DENF individually, which is important for potential decision-making. For example, identifying the effects of individual *Aedes* habitats (e.g. tire shops and trees) can provide more intuitive results than grouping them in a PCA. As a result five variables were selected for subsequent modeling with a VIF value < 3: population density, tree density, relative proximity to rivers, relative proximity to tire shops, and relative proximity to plant nurseries.

The PCA’s purpose is to reduce and simplify the variables into new variable (components) that explain a large degree of variation without collinearity between the components. Tables 2 and 3 provide the results of the PCA analysis and include the variables that were included – which were not included in the VIF testing. Table 2 describes the variance explained by the top three principal components and Table 3 shows the variables that belong to each component. After examining the eigenvalues and component loadings with coefficients > 0.40, the first two components were selected for inclusion in the space-time modeling, which explains 60.8% of the variance (42% and 18.8%, respectively).

**Table 2.** Explained variance by top three principal components

| Component | Extraction Sums of Squared Loadings |  |  | Rotation Sums of Squared Loadings |  |
| --- | --- | --- | --- | --- | --- |
|  | Total | Variance | Cumulative % | Total | % of Variance |
| 1 | 4.516 | 45.163 | 45.163 | 4.203 | 42.032 |
| 2 | 1.703 | 17.031 | 62.194 | 1.882 | 18.817 |
| 3 | 1.072 | 10.719 | 72.912 | 1.206 | 12.064 |

**Table 3.** Rotated component loadings with coefficient > 0.40

| Variable | Component |  |  |
| --- | --- | --- | --- |
|  | 1 | 2 | 3 |
| Strata* | .856 |  |  |
| % OHH** |  | .749 |  |
| % HW** |  | .657 |  |
| % Work** | .784 |  |  |
| % Retired** | .781 |  |  |
| % Disabled** |  |  | .782 |
| % Student** |  | .873 |  |
| % Married** | .941 |  |  |
| % EmptyHH** | .484 |  | -.514 |
| % HighEduc** | .940 |  |  |

\*Neighborhood strata classification between 1 and 6.
\*\*proportion (%) of individuals in each category: (1) Occupied Households (2) Work from Home (3) Retired persons (4) Disabled persons (5) Students (6) Married persons (7) Empty Households (8) holds college degree or higher

PC1 is strongly associated with (correlation close to 0.5 and above) neighborhood strata, individuals who are employed, retired persons, married individuals, empty households, and individuals with high education. PC2 mainly explains occupied households, individuals who do housework, and individuals who are students. PC3 explains the combined effect from disabled individuals but was excluded from further analysis and it was included as a separate variable in further VIF (variance inflation factor) testing with the remaining covariates. PC1 can be interpreted as employed, higher-income people who are likely to be older and married due to the large coefficients of retired and married individuals. PC2 can be interpreted as people who are likely to spend more time at home than those in PC1 because they are either students or do housework for a living. PC2 probably also captures younger individuals due to the inclusion of students.

#### 2.2.1 Selecting Lagged Weather Variables

First, VIF testing was used to assess multicollinearity among the candidate weekly weather variables (Table 1) and remove the most inter-correlated variables while retaining the most independent. Seven variables (with VIF values < 3) were selected for the subsequent cross-correlation analysis: Tavg, DTRmax, RHrng, RainT, RainD, CoolD, and WarmD. Next, lagged cross-correlations were computed between weekly DENF rates and each weather variable over an 8-week window (Table 4). Such time frame was considered the most biologically plausible based on existing literature of how weather affects the full vector lifecycle and viral transmission dynamics^20,34^ (see Table 5). Finally, an optimal lag for each weather variable was selected among those weeks exhibiting a statistically significant (at the 5% level; adjusted for multiple testing) cross-correlation within the 1-8 week lag window (0-week lags were not considered due to their lack of predictive potential). For six variables (Tavg, DTRmax, RainT, RainD, CoolD, WarmD), the selected optimal lags (5-weeks, 5-weeks, 3-weeks, 5-weeks, 2-weeks, 5-weeks, respectively) exhibited the maximum cross-correlation coefficient (Table 4). For RHrng, the maximum correlation occurred at 6-weeks, but the 3-week lag was deemed optimal since existing literature suggests that relative humidity is most influential on adult vectors and the gonotrophic cycle (Table 5). The optimal lags were included in the ST-CAR models.

**Table 4.** Lagged cross-correlations between DENF rates and weekly weather variables during 2015-2016 in Cali, Colombia

2015-2016 in Cali, Colombia
| Lag<br>(Weeks) | Tavg | RainT | RHrng | DTRmax | RainD | CoolD | WarmD |
| --- | --- | --- | --- | --- | --- | --- | --- |
| 0 | 0.419* | -0.207 | -0.012 | 0.463* | -0.508* | 0.202 | 0.507* |
| -1 | 0.285* | -0.413* | -0.057 | 0.150 | 0.011 | -0.109 | 0.261* |
| -2 | 0.032 | 0.098 | 0.212 | 0.077 | 0.337* | -0.354** | 0.095 |
| -3 | 0.074 | -0.458** | -0.448** | 0.029 | -0.012 | -0.166 | -0.012 |
| -4 | 0.257* | -0.092 | -0.166 | 0.105 | -0.488* | 0.081 | 0.201 |
| -5 | 0.369** | -0.262* | -0.362* | 0.236** | -0.542** | 0.201 | 0.287** |
| -6 | 0.255* | -0.316* | -0.557* | -0.104 | -0.359* | 0.304* | 0.014 |
| -7 | 0.221 | -0.011 | -0.473* | 0.079 | -0.338* | 0.283* | -0.047 |
| -8 | 0.242 | -0.315* | -0.098 | -0.088 | -0.309* | 0.181 | -0.156 |

\*Significant at the 5% level (after adjustment via a Bonferroni correction for multiple testing)
\*\*Selected Lag for ST-CAR modeling

**Table 5.** Temporally-lagged weather variables & correlation with *Aedes*’ life cycle (*Based on 20, 34); #Total lag (weeks prior to lab confirmation) - accumulated range for each stage in the vector-dengue transmission cycle

| Development<br>Stage | Range<br>(Days)<br>* | Total Lag<br>(Weeks) # | Variable | Expected<br>Relation | Rationale | Primary<br>Sources |
| --- | --- | --- | --- | --- | --- | --- |
| **Larval/Pupa Development**<br>(vector grows in water) | 10-21 | **3-8** | RainT | Positive | More total rain produces more stagnant pools; promotes larval development | 38, 39 |
|  |  |  | RainD | Positive | More frequent rain produces more stagnant pools; promotes larval development | 40, 41 |
|  |  |  | Tavg | Positive | Warmer temperatures promote larval development | 42, 43 |
|  |  |  | DTRmax | Negative | Smaller ranges imply fewer hours with cold temperatures; greater larval survival | 20, 44 |
|  |  |  | RHrng | Negative | Smaller ranges imply fewer dry days; stagnant pools quickly evaporate | 20, 45 |
|  |  |  | WarmD | Positive | More warmer days promote greater larval densities | 44, 46 |
|  |  |  | CoolD | Negative | Fewer cold days promote greater larval survival | 44, 46 |
| **Gonotrophic Cycle**<br>(vectors feed on humans) | 3-7 | **2-5** | RainT | Negative | Rainfall tends to coincide with cooler temperatures; less vector feeding | 20, 47 |
|  |  |  | RainD | Negative | Rainfall tends to coincide with cooler temperatures; less vector feeding | 20, 47 |
|  |  |  | Tavg | Positive | Vector feeding more frequent at warmer temperatures | 48, 49 |
|  |  |  | DTRmax | Negative | Vector feeding more frequent at warmer temperatures; smaller DTRmax implies fewer cold hours | 20, 42 |
|  |  |  | RHrng | Positive | Vector feeding more frequent during dry periods; larger RHrng implies more dry days | 20, 45 |
|  |  |  | WarmD | Positive | Vector feeding more frequent at warmer temperatures | 50 |
|  |  |  | CoolD | Negative | Vector feeding more frequent at warmer temperatures; fewer cold hours | 50 |
| **Extrinsic Incubation**<br>(virus matures in vector) | 7-15 | **1-4** | RainT | Unknown | No known link between rainfall and extrinsic incubation | N/A |
|  |  |  | RainD | Unknown | No known link between rainfall and extrinsic incubation | N/A |
|  |  |  | Tavg | Positive | Viral development more rapid at warmer temperatures | 48, 51 |
|  |  |  | DTRmax | Negative | Viral development more rapid at warmer temperatures; smaller DTRmax implies fewer cold hours | 50 |
|  |  |  | RHrng | Positive | Viral development more rapid in humid environments; smaller RHrng implies fewer dry days | 52, 53 |
|  |  |  | WarmD | Positive | Viral development more rapid at warmer temperatures | 50 |
|  |  |  | CoolD | Negative | Viral development more rapid at warmer temperatures; fewer cold hours | 50 |
| **Intrinsic Incubation**<br>(virus matures in humans) | 1-12 | **0-2** | N/A | N/A | No known link between weather and intrinsic incubation | N/A |
| **Dengue Reported**<br>(lab confirmation) | --- | --- | N/A | N/A | N/A | N/A |
| **Total** | 21-55 | **0-8** |  |  |  |  |

#### 2.2.3 Poisson GLMs

First, a Poisson generalized linear model (GLM) is computed to detect significant effects of independent variables on a dependent variable (DENF risk); and the presence of spatiotemporal autocorrelation in the residuals. The Poisson GLM is defined as:

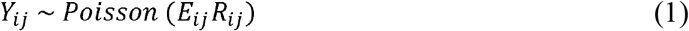

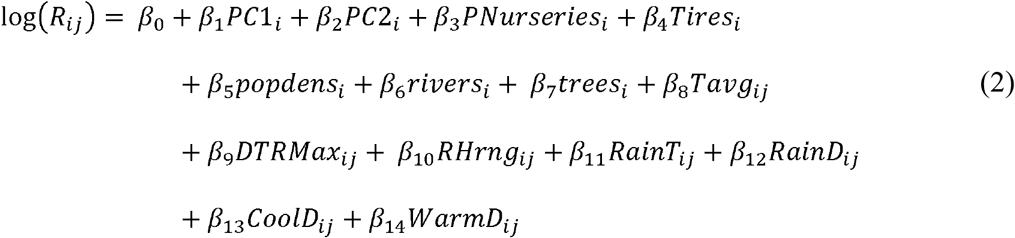

where *Y*_*ij*_ is the observed DENF count in neighborhood *i* at week *j*; *E*_*ij*_ is the expected DENF count in neighborhood *i* at week *j*; and *R*_*ij*_ is the disease risk in neighborhood *i* at week *j* (see supplemental materials for the results of the Poisson GLM).

Global Moran’s I^54^ was then computed to detect spatial autocorrelation of the Poisson GLM residuals for each time period. Essentially, the Global Moran’s I test determines if there is evidence of unexplained spatial autocorrelation in the residuals; and if positive spatial autocorrelation is detected, then the assumption of independence is not valid for the data, and spatiotemporal autocorrelation should be considered when estimating covariate effects on the dependent variable. The global Moran’s I index ranges from −1 to 1, while −1 indicates strong negative spatial autocorrelation, 0 indicates complete spatial randomness, and 1 indicates strong positive spatial autocorrelation; and the statistic is defined as:

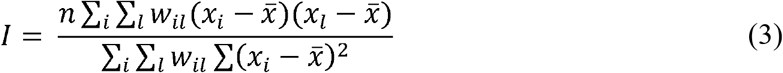

where *n* is the total number of neighborhoods, *w*_*il*_ is the spatial weight between neighborhood *I* and *l*, 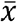 is the mean of residuals for all neighborhoods, *x*_*i*_is the residual value in neighborhood *i*, and *x*_*l*_is the residual in neighborhood *l*. The Moran’s I tests were conducted in RStudio 1.2.5 with R version 3.6.

#### 2.2.4 ST-CAR Modeling

Next, a Bayesian hierarchical model^24,25^ is defined using a Poisson data model (for case/population data). The model “represents the spatio-temporal pattern in the mean response with a single set of spatially and temporally autocorrelated random effects. The effects follow a multivariate autoregressive process of order 1^54^. In other words, when going from one week to another (e.g. *j* +1), it will yield an effect on the dependent variable (DENF risk). Therefore, this model examines linear trends which can be interpreted as how DENF risk is influenced across time.

It is assumed that the estimated effect on DENF risk in the ST-CAR model is not specific to a particular week, but a process that is influenced by the covariate data across the weeks (temporal unit). Suppose a study region is divided into a collection of *N* non-overlapping areal units (e.g. neighborhoods) indexed by *i* ∈ {1….,N}; and the data is observed for multiple time periods, that is: *j* ∈ {1….,T}. As suggested before, ST-CAR models utilize prior distributions, where the CAR distributions state that adjacent variables in space or time are conditionally autocorrelated, and non-adjacent variables are conditionally independent. The spatial weight matrix is defined as: *W = (w*_*ik*_, where a value of 1 indicates that *i* and *k* are spatially adjacent, and 0 otherwise. Since it is unknown where a person was infected, the abovementioned adjacency matrix is a proxy for *Aedes’* maximum range – since they do not fly more than 400 meters from where they emerged as larvae^10^. A temporal weight matrix can also be defined as *D* = (*d*_*jt*_), where a value of 1 is given if *t* – *j* = 1, and 0 otherwise. The first part of the model is defined as:

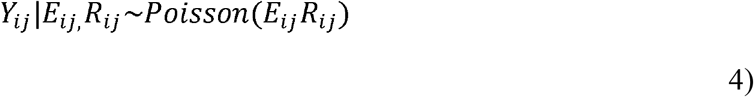

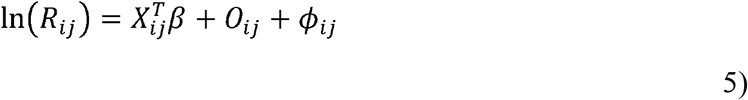

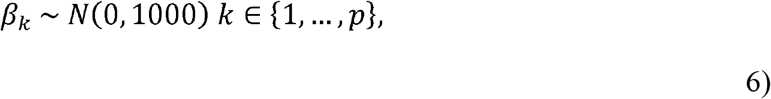

where *Y*_*ij*_ is the observed DENF count in neighborhood *i* at week *j*; *E*_*ij*_ is the expected disease count in neighborhood *i* at week *j*; and *R*_*ij*_ is the disease risk in neighborhood *i* at week *j*. 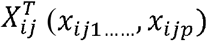 is a vector of known covariates *p* for neighborhood *i* and week *j*. The parameter β is an associated *p* x 1 vector of regression parameters, which can come from the initial Poisson GLM in Equations 1 and 2. The term *o* is a vector of known offsets (*o*_1_,….., *o*, where *o*_*j*_ is a *K* * 1 column vector of offsets (expected DENF cases) for week *j* (*o*_*1j*_,… …, *o*_*kj*_. An offset variable is used to scale the modeling of the mean in Poisson regression with a log link, which is the case in the above model. For example, since the dependent variable is rates, the offset can enforce that 10 cases of DENF in one week is not the same magnitude as 10 cases of DENF in 6 weeks. The parameter *ϕ*_*ij*_ denotes spatiotemporally autocorrelated random effects for neighborhood *i* and week *j*. A variety of spatiotemporal structures can be fit for *ϕ*_*ij*_. Here, we use a model that estimates the evolution of the spatial response surface over time without forcing it to be the same for each time period (Rushworth et al. 2014).

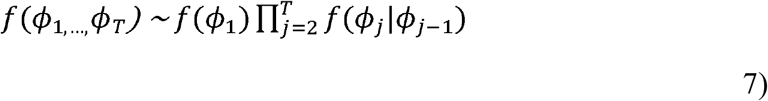

Where 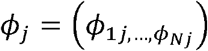 is a vector of random effects for week *j*. Temporal autocorrelation is enforced because *ϕ*_j_ depends on *ϕ*_*j-1*_. *f ϕ*_1_ enforces spatial autocorrelation in the random effects, where the spatial structure is defined in the CAR prior in Equation 8:

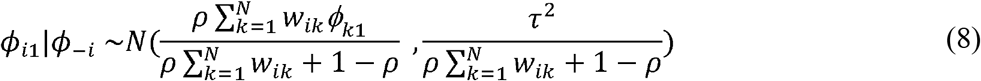

Where *ρ* controls the spatial autocorrelation with *ρ* = 1 indicating strong spatial autocorrelation, which is conditional upon the mean random effects of adjacent neighborhoods. *ρ*= 0 represents independent random effects with a constant mean and constant variance. The conditional precision is controlled by, where precision is higher when more prior information (e.g. adjacent neighborhoods) is borrowed to determine the posterior estimates. Equation 9 (CAR Prior) enforces temporal autocorrelation in the random effects and is defined as:

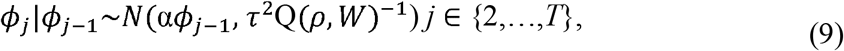

Where Q(*ρ, W*) is a precision matrix, that is defined as *ρ* (diagonal(W1) – W) + (1 – *ρ*)*I*, where *I* is a N x N identity matrix and the ‘1’ is a vector of ones (N x 1). The α controls the temporal autocorrelation, where 0 is temporally independent and 1 is strong temporal dependence. The CAR priors also include weakly informative hyperpriors (i.e. probability distribution from priors to inform/update posterior values), which are the three parameters defined below:

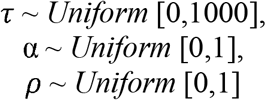

The values of the hyperpriors are selected in a way so that our Bayesian inferences are robust and not sensitive to these choices. For example, a non-stationary spatial process would occur when *ρ* = 1; and non-stationary temporal process would occur if α = 1. Overall, the ST-CAR model states that when going from one week to another (*j* + 1), it yields an effect on the dependent variable (DENF risk), which is influenced by spatially and temporally dependent covariates. In other words, DENF risk in a target neighborhood is influenced by current and past values of DENF risk and covariates at surrounding neighborhoods and time periods (which is a process that evolves over time). Conceptually, a spatial example would suggest that a neighborhood with low risk of DENF would have an increased risk of DENF if an adjacent neighborhood reported a high risk of DENF (dependence/autocorrelation).

Statistical inference is derived from Markov Chain Monte Carlo (MCMC) simulations^56^. MCMC is a popular technique for Bayesian inference when posterior distributions are not available in closed forms. We used MCMC to generate samples from the posterior distributions induced by our ST-CAR models and used them for parameter and density estimation. For this study, we selected 220,000 MCMC samples; initial 20,000 samples are removed as burn-in; and the thinning parameter is set to 10 (keeping every 10^th^ value and removing all others), which thins the samples to reduce autocorrelation of the Markov Chain. As a result, 20,000 samples are used for statistical inference. The deviance information criterion (DIC) was used for model assessment and comparison, where lower values of DIC indicate a better model fit. The results of two models are presented in section 3: Model 1 (DENF with no lags); Model 2 (DENF with lagged weather variables). Using CARBayesST^24^ in R, the two Poisson GLM models (DENF with no lags; DENF with lags) were each fitted to the ST-CAR model described above.

## 3. Results

### 3.1 ST-CAR Results

The following subsections contain summaries of model results; relative risk estimates for each independent variable; and mapping the posterior estimates of DENF rates in Cali for particular time periods (temporal cross-sections).

#### 3.1.1 Model 1

Table 6 (left) summarizes the results of Model 1 (DENF with no lags). The 95% credible intervals do not contain a value of 0, indicating that all the covariates exhibit influential relationships with DENF risk at the neighborhood level in Cali between January 2015 and December 2016. The spatial autocorrelation value of 0.98 indicates that there is very strong spatial dependence of the data after adjusting for covariate effects. The temporal autocorrelation value of 0.11 suggests that there is some presence of temporal dependence of the data after adjusting for covariate effects. Therefore, DENF risk in Cali at the neighborhood level is influenced by DENF rates and covariates in surrounding neighborhoods and time periods (weeks). Table 6 (left) also shows the relative risk estimates (%) of DENF in Cali between January 2015 and December 2016. The results suggest a 4.7% increase in DENF risk for PC1; a 4.7% decrease for PC2; a 12.9% increase for proximity to plant nurseries; a 5.5% increase for proximity to tire shops; a 1.5% decrease for population density; a 11.6% decrease for proximity to rivers/ravines; a 36% increase for tree density; a 635% increase for average temperature; a 26.6% increase for days with maximum temp. > 32 °C; a 139% increase for relative humidity range; a 45.8% decrease for total rainfall; a 16.8% decrease for total rain days; a 76.4% increase for cool days; and a 73.6% decrease for warm days when adjusted for other variables. The results will be further explained in the discussion.

**Table 6:** ST-CAR Model Results (Model 1 – no lags); (Model 2 – lagged weather variables)

|  | Model 1 – DIC: 67,055.98 |  |  |  | Model 2 – DIC: 66,954.16 |  |  |  |
| --- | --- | --- | --- | --- | --- | --- | --- | --- |
|  | 2.50% | Median | 97.50% | RR (%) | 2.50% | Median | 97.50% | RR (%) |
| Intercept | -2.0296 | -1.6427 | -1.3183 | NA | -0.8195 | -0.5423 | -0.3055 | NA |
| PC1 | 0.0159 | 0.0463 | 0.0785 | 4.7 | 0.0459 | 0.0732 | 0.1 | 7.6 |
| PC2 | -0.0885 | -0.0475 | -0.0068 | -4.6 | -0.0042 | 0.0382 | 0.0798 | 3.9 |
| Pnurseries | 0.0203 | 0.1213 | 0.2215 | 12.9 | 0.0841 | 0.1806 | 0.2787 | 19.8 |
| Tires | -0.0838 | 0.0531 | 0.1902 | 5.5 | -0.023 | 0.1118 | 0.2469 | 11.8 |
| Popdens | -0.173 | -0.0146 | 0.1482 | -1.5 | -0.3296 | -0.1749 | -0.0176 | -16 |
| Rivers | -0.2473 | -0.1233 | -0.0029 | -11.6 | -0.2532 | -0.1381 | -0.0227 | -12.9 |
| Trees | 0.1254 | 0.3071 | 0.4909 | 36 | 0.0901 | 0.2626 | 0.4404 | 30 |
| Tavg(L5) | 1.5864 | 1.9947 | 2.3546 | 635 | -0.6022 | -0.2908 | 0.0415 | -25.2 |
| DTRMax(L4) | -0.24 | 0.2361 | 0.9014 | 26.6 | 0.4579 | 0.8115 | 1.2709 | 125.1 |
| RelHRng(L3) | 0.4641 | 0.8713 | 1.342 | 139 | 0.4317 | 0.615 | 0.8213 | 85 |
| RainT(L3) | -1.3716 | -0.613 | 0.003 | -45.8 | -2.8222 | -2.4343 | -2.0671 | -91.2 |
| RainD(L5) | -0.5249 | -0.1837 | 0.1929 | -16.8 | -0.9178 | -0.7063 | -0.4845 | -50.7 |
| CoolD(L2) | 0.0013 | 0.5678 | 1.1938 | 76.4 | -0.8337 | -0.2914 | 0.1285 | -25.3 |
| WarmD(L5) | -1.7085 | -1.3392 | -0.9888 | -73.8 | 0.2861 | 0.6383 | 0.9196 | 89.3 |
| $\rho$ | 0.977 | **0.9829** | 0.9872 | NA | 0.977 | **0.9824** | 0.9867 | NA |
| $\alpha$ | 0.0736 | **0.1189** | 0.1636 | NA | 0.0737 | **0.1193** | 0.1651 | NA |

#### 3.1.2 Model 2

Table 6 (right) summarizes the results of Model 2. The 95% credible intervals do not contain a value of 0, indicating that all the covariates exhibit influential relationships with DENF risk at the neighborhood level in Cali between January 2015 and December 2016. The spatial autocorrelation value of 0.98 indicates that there is very strong spatial dependence of the data after adjusting for covariate effects. The temporal autocorrelation value of 0.11 suggests that there is some presence of temporal dependence of the data after adjusting for covariate effects. Therefore, DENF risk in Cali at the neighborhood level is influenced by DENF rates and covariates in surrounding neighborhoods and time periods (weeks). The DIC (66,964.16) is slightly lower than Model 1 (DENF with no lags – DIC = 67,055.98). In general, the lagged weather variables in Model 2 shrunk the confidence intervals of the coefficients (most notably for average temperature and Days Temp Max); which decreases the uncertainty of our model’s risk estimates.

Table 6 (right) also shows relative risk estimates of DENF in Cali between January 2015 and December 2016. The results suggest a 7.6% increase in DENF risk for PC1; a 3.9% increase for PC2 (negative relationship [decreased risk] in Model 1); a 19.8% increase for proximity to plant nurseries; a 11.8% increase for proximity to tire shops; a −16% decrease for population density; a 12.9% decrease for proximity to rivers/ravines; a 30% increase for tree density; a 25.2% decrease for average temperature; a 125.1% increase for days with maximum temp. > 32 °C; an 85% increase for relative humidity range; a 91.2% decrease for total rainfall; a 50.7% decrease for total rain days; a 25.3% decrease for cool days; and a 89.3% increase for warm days. The negative to positive relationship between DENF and PC2 observed when comparing

Models 1 and 2 is difficult to interpret. This could be due to the lagged weather variables affecting the posterior estimates. The magnitude of PC2 (low RR) is much lower than the other independent variables, therefore, we hypothesize that people that spend more time at home (PC2) may or may not be more susceptible to DENF and further investigation is required.

#### 3.1.1 Mapping the posterior estimates of DENF

Figure 3 provides maps of the temporal cross-sections of Model 2 posterior values for each neighborhood of DENF rates (per, 1,000) in Cali between 2015 and 2016. The weekly estimates were aggregated by month for visualization purposes – January 2015, July 2015, December 2015, January 2016, July 2016, and December 2016, respectively.

**Figure 3.**
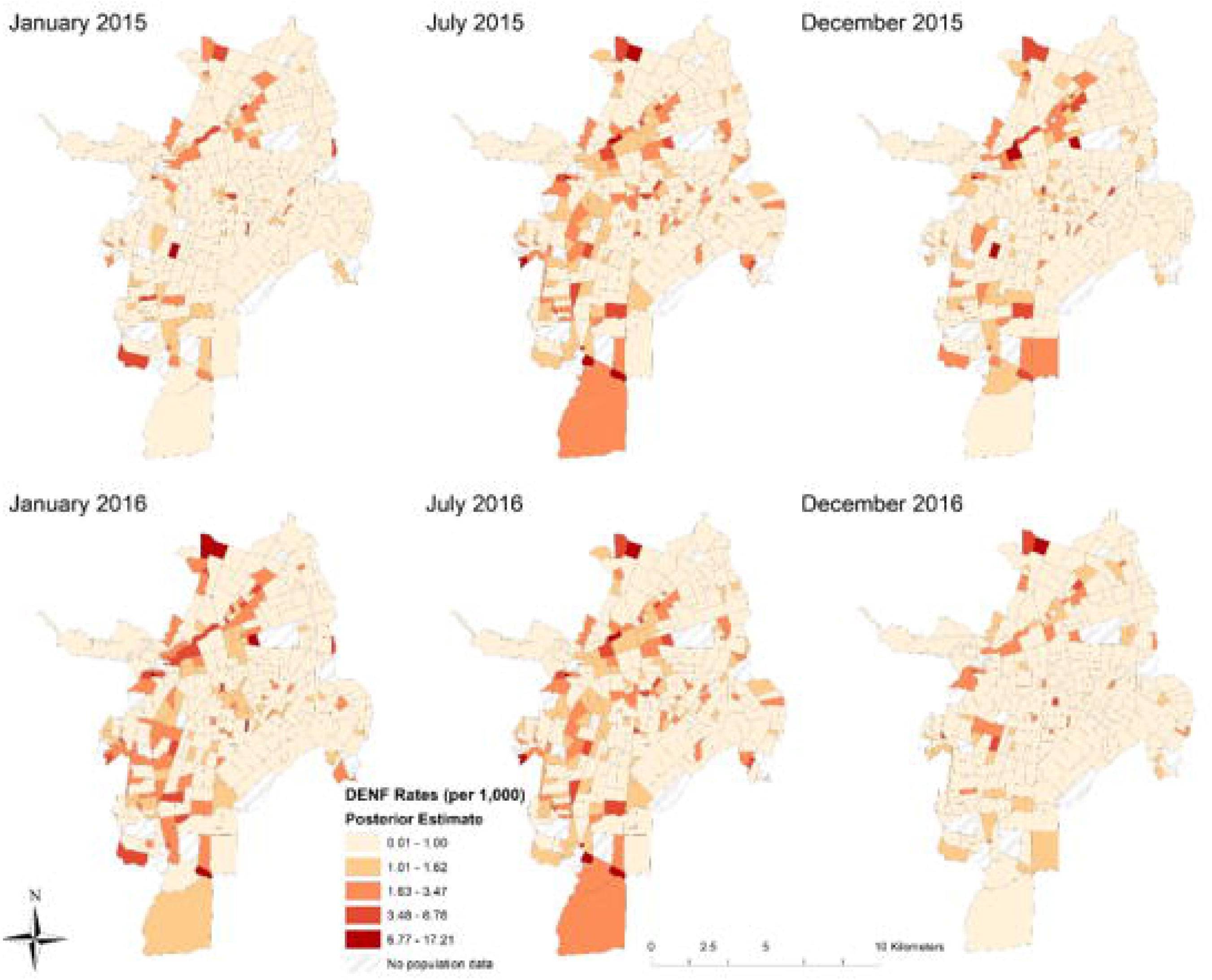
Temporal cross-sections of model 2 posterior values for each neighborhood of DENF rate (per 1,000) in Cali.

When comparing the six temporal cross-sections, July 2016 experienced the highest DENF risk (estimated posterior mean values) after accounting for the 14 covariates (including the lagged weather variables). Interestingly, some locations with high rates of DENF are classified as middle (3 or 4) or high strata (5 or 6). These middle and high strata neighborhoods are adjacent to low strata neighborhoods (1 or 2), which suggests that there is spatial-temporal dependence between them. In other words, there is evidence that middle and high strata neighborhoods are at higher risk when surrounded by lower strata neighborhoods after accounting for the covariates in the models. Some of the highest proportion of cases were observed in the eastern portions of Cali (now shown here), but also include some of the highest and most densely populated neighborhoods of the city. After computing rates per 1,000 persons (Figure 3), the eastern portions of Cali (which include some of the poorest neighborhoods) have lower reported rates of DENF than central, southern, and western regions of Cali.

## 4. Discussion

This study is the first of its kind to model space-time risk at the neighborhood- and weekly-levels of DENF across two years of disease surveillance data; while also incorporating temporally lagged weather variables in a ST-CAR approach. Coupling the lagged weather variables with the spatial covariates of DENF risk, the models include neighborhood-level effects explaining where and why certain locations are more at-risk than others. A quintessential spatiotemporal model decomposes the variability in the outcome in large scale variations (the mean function) and small-scale variation (the spatiotemporal random effects)^22^. Independent variables were used to characterize large-scale variations, while spatiotemporal autocorrelation was used to characterize small-scale variations. The lagged independent variables modified the large-scale variations and for this data it seems they have no effect on small scale variations. The predictive power of the models remains the same, very close DIC values (see Table 6). That is why we are seeing quite robust estimates of spatial autocorrelation parameters in both models. There are many key findings that warrant further investigation and explanation.

### 4.1 Influence of Socioeconomic and Environmental Factors

First, there is very strong evidence that there is both spatial and temporal dependence between DENF risk and the significant covariates for adjacent neighborhoods in Cali. Although DENF had a much lower temporal autocorrelation (value of 0.11 for both Models 1 and 2), this can be explained by the distribution of cases between 2015 and 2016 – there were three distinct peaks, but DENF remains a persistent threat due to the four serotypes of the virus. The very strong spatial autocorrelation values (although extremely high) for DENF suggest that outbreaks in adjacent neighborhoods are strongly related (i.e. living next to a neighborhood with high cases will strongly influence DENF risk and cases in your neighborhood of residence).

When examining the results of the three socioeconomic covariates (PC1, PC2, and population density), the increased risk of DENF for PC1 is an interesting finding. This corroborates with Delmelle et al. (2016)^30^, who suggest that in the southern part of Cali, houses are typically bigger with relatively larger yards, which may provide a suitable habitat for *Aedes* to breed. Neighborhoods with a high proportion of people in the PC1 category (e.g. employed, older, more educated) are also typically adjacent to middle or lower strata neighborhoods (with the exception of the extreme South), which also may increase the risk of disease due to the very strong evidence of space-time dependence between the locations, as suggested by the models. An increased risk of DENF was reported for neighborhoods with higher proportion of individuals in the PC2 category (i.e. work from home and students). This finding corroborates with strong evidence that *Aedes* proliferates in and around homes^57,58,59^; and it has also been found that cases can substantially decline if *Aedes* trap interventions are put in place in at-risk communities^60^.

Another unexpected result was the negative relationship between population density and DENF. One explanation can be that some of the densely populated neighborhoods in the eastern part of Cali have a high concentration of Afro-Colombian population, which has been suggested to be less susceptible to the viruses^61,62^. Furthermore, there is evidence that shows that low density areas with poor infrastructure may have increased *Aedes* presence^63^ and DENF’s complex immunology and the herd immunity resulting after infection from the viruses^64^ may have contributed to these patterns. Amongst other factors, high density of populations may not necessarily be a main risk factor of VBD transmission^65^. While increases in population and urbanization will undoubtedly increase risk of VBD transmission, the true effect of population density may vary at fine spatial levels (e.g. neighborhoods). Public health authorities typically carry out fumigation and education programs in neighborhoods with higher population densities, which may result in a lack of targeted interventions in less dense areas with *Aedes* populations (e.g. sewers, green areas, etc.). Vector control efforts in Cali include storm sewer control, spraying, use of guppy fish, visits to health facilities and homes, and community educational campaigns^35^.

Closer proximity to plant nurseries, tire shops, and higher tree density all exhibited an increased risk of DENF. Plant nurseries and tire shops are common breeding grounds of *Aedes*, thus neighborhoods within close proximity are generally at a higher risk of disease transmission. Tree density may also be a significant risk factor of *Aedes* presence since studies have shown that high tree shade density stimulates breeding; and tree holes are suitable water containers where *Aedes* have been found in abundance^67,68^.

Closer proximity to rivers and ravines (i.e. any moving bodies of water) resulted in significantly lower risk of DENF. *Aedes* require stagnant water as a breeding ground, therefore, the flowing water of a river or ravine would prove to be an unsuitable habitat for the mosquitoes. Although flooding events during the rainy season could create stagnant water sources surrounding the rivers, however, floods may also act as a disruptive force on *Aedes* habitat by flushing out their breeding sites and eggs^69,70^. Further research using remote sensing techniques (such as flow analysis) could provide insight to areas prone to stagnant water.

### 4.2 Influence of Weather Factors

In general, the use of optimally lagged weather variables shrunk the confidence intervals about the mean model coefficients and relative risk estimates. Specifically, the individual relationships between each lagged weather covariate and disease risk became either consistently positive or consistently negative (except for average temperature). Moreover, since the optimal lags were different among the weather covariates, our results imply that individual covariates impact different stages of the vector lifecycle (see Table 5). Given that such relationships could inform future space-time modeling and mitigation efforts, the most likely physical connections between local weather conditions and the *Aedes* lifecycle (Tables 4 and 5) are discussed in greater detail based on our Model-2 results (Table 6).

Variables with an optimal 5-week lag (Tavg, DTRmax, RainD, WarmD) are most influential on larval development. Both DTRmax and WarmD exhibit the expected positive relationship (Table 5) and large predictive importance based on relative risk estimations (Table 6), while RainD exhibits the expected negative relationship with moderate importance. Despite Tavg having an unexpected negative relationship with DENF, its predictive importance is relatively small. Five weeks before above-average DENF rates, the weather is often characterized by multiple days with short-lived rain showers (RainD is above average, its correlation with DENF is negative, and the regression coefficient is negative), yet each day experiences sufficient sunshine to allow the daily maximum temperature to exceed 32°C (WarmD is above average, its correlation with DENF is positive, and the regression coefficient is positive).

The short-lived rain showers also induce evaporative cooling that lowers the daily minimum temperature and increases the daily temperature range (DTRmax is above average, its correlation with DENF is positive, and the regression coefficient is positive), leading to slightly cooler, but above-average daily mean temperatures (Tavg remains above average, its correlation with DENF remains positive, but the regression coefficient is slightly negative and the relative risk is small). Overall, regular rainfall combined with average to above-average temperatures produce numerous stagnant pools within a favorable thermal environment for prolific larval development.

Variables with an optimal 3-week lag (RainT and RHrng) are most influential on the gonotrophic cycle. Both RainT and RHrng exhibit the expected relationships (Table 5) with similar moderate levels of predictive importance (Table 6). Weather conditions three weeks prior to above-average DENF rates are often characterized by minimal rainfall (RainT is below average, its correlation with DENF is negative, and the regression coefficient is negative) and clear skies, which allows the relative humidity to fluctuate between small daytime values and large nighttime values (RHrng is above average, its correlation with DENF is positive, and the regression coefficient is positive). Overall, the relatively dry conditions maximize solar heating, minimize evaporative cooling, and promote the warm temperatures most favorable for Aedes feeding.

Finally, variables with an optimal 2-week lag (CoolD) are most influential on the gonotrophic cycle and extrinsic incubation. Specifically, CoolD exhibits the expected negative relationship (Table 5), but its relative importance is small and roughly equivalent to Tavg. Weather conditions two weeks before above-average DENF rates often exhibit above-average temperatures (CoolD is below average, its correlation with DENF is negative, and the regression coefficient is negative); whereby, the warmer temperatures will accelerate both vector feeding and viral replication within the vector, which increases the potential for transmission to humans.

Overall, such consistent multi-lag relationships reinforce the idea that DENF outbreak dynamics are dependent on a complex combination of weather conditions 2-5 weeks prior (Eastin et al. 2014). Careful monitoring of weather conditions within this time window could optimally inform any sub-seasonal DENF mitigation efforts. Moreover, it should be noted that a moderate El Niño occurred during our 2015-2016 study period^71^ and there is strong evidence that major DENF epidemics are more severe during El Nino events^20,72^. Therefore, any sub-seasonal mitigation efforts should also account for inter-seasonal climate variability.

### 4.3 Limitations

Despite the strengths and contributions of this research, there are notable limitations and areas of future work that is worth discussing. First, the underreporting of cases and unmatched addresses during the geocoding process likely undermines the true burden of DENF. Underreporting is also a major issue in the low strata neighborhoods and also not uniform across the city, while surveillance systems can improve the identification of risk factors throughout the city^73^. Second, the socioeconomic and demographic data was a mix from the Colombian National Census and 2010 population estimates. Colombia recently administered a new national census (the first since 2005) but is currently unavailable. Using 2005 and 2010 data for this study will bias the results, but the neighborhood classifications (strata) mostly remained unchanged. The uncertainty resulting from using outdated census data is a common limitation found in many studies in Latin America and developing countries. Several articles examining patterns of dengue fever in Colombia have recognized outdated population census being a critical issue that could affect the findings of their research^74-79^. Until more recent census are available, different population growth models could address this issue, or relying on disaggregated population projection, using dasymetric mapping^80,81^. Third, including vector surveillance data (presence/absence) in each neighborhood would improve the accuracy of the relative risk estimates^17^. Fourth, the spatial weight matrix only considered adjacent neighborhoods as “neighbors”, we recognize that individual activity spaces expand far beyond locations nearby their home. Future research can implement different spatial and temporal weight matrices for sensitivity analysis purposes. Fifth, the weather conditions and severity of outbreaks for DENF varied between 2015 and 2016, which may have affected the model results. Future work can disaggregate the years and run two separate models for further examination. Sixth, the space-time patterns of the DENF outbreaks did not exhibit much seasonality, which is likely due to only using two years of data. Further work can utilize 5-10 years of DENF and weather data to detect potential seasonal patterns of the epidemics. Finally, weather variability is represented by a single weather station (and thus limited to the temporal domain). Future work would benefit from multiple weather stations that can document spatial variability across the region.

### 4.4 Opportunities

Chikungunya (CHIK) and Zika are transmitted by the same vector (*Aedes*), and further investigation can develop a multivariate space-time CAR model to examine which neighborhoods are at higher risk for one disease, two diseases, or all three concurrently. Multivariate space-time (MVST) approach in CAR modeling is still in its infancy stages^24^ and the development of such models can more accurately examine and compare the co-occurrence of diseases transmitted by the same vector. A long-term goal is to develop a multi-parameter early warning system (EWS) that informs public health officials and motivates effective vector surveillance and control measures. The space-time models and methods described herein represent progress toward that goal. However, effective EWS development would require larger and more comprehensive databases (for both robust model development and independent validation) than those currently available. As noted above, an EWS system would benefit from longitudinal information regarding socioeconomic, demographic, and environmental variables combined with spatial information regarding weather variability across the region.

## 5. Conclusion

A ST-CAR modeling approach was utilized to examine significant socioeconomic, demographic, environmental, and meteorological risk variable of DENF in Cali, Colombia during 2015 and 2016. The temporally lagged weather covariates can significantly estimate when risk of transmission is highest, and the spatial covariates can help explain the differences in disease risk at the neighborhood-level. Adding weather and climate data to a space-time model can improve disease surveillance, especially for VBDs that require specific conditions for transmission to occur. This study demonstrated that there was strong spatial and temporal dependence between adjacent neighborhoods and time periods, which provides strong evidence that DENF transmission are influenced by characteristics and phenomena occurring in surrounding locations. We also provide evidence that DENF is not just a disease of the poor; although risk factors may be higher in neighborhoods of lower socioeconomic status, we have shown that the transmission dynamics of DENF are place- and temporally based. Despite this study being retrospective in nature, the modeling approach can be applied in a contemporary surveillance setting when significant outbreaks have not yet occurred, highlighting at-risk areas to help promote proactive community health, improve public health educational campaigns, targeted interventions. We hope that this research influences further small area space-time analysis, since we support the notion that disease prevention (in general) should start at the neighborhood and community level.

## Data Availability

The data is currently unavailable due to prior data release agreements.

## Acknowledgements

The authors would like to thank the editor and anonymous reviewers for their constructive feedback that ultimately improved the quality of this paper.

